# Estimated incidence rate of specific cardiovascular and respiratory hospitalizations attributable to Respiratory Syncytial Virus among adults in Germany between 2015 and 2019

**DOI:** 10.1101/2024.07.19.24310503

**Authors:** Caihua Liang, Aleksandra Polkowska-Kramek, Caroline Lade, Lea Johanna Bayer, Robin Bruyndonckx, Bennet Huebbe, Worku Biyadgie Ewnetu, Pimnara Peerawaranun, Maribel Casas, Thao Mai Phuong Tran, Gordon Brestrich, Christof von Eiff, Bradford D. Gessner, Elizabeth Begier, Gernot Rohde

**Affiliations:** Pfizer Inc-New York (USA); P95 Epidemiology & Pharmacovigilance, Leuven, Belgium; Pfizer Pharma GmbH - Berlin (Germany); Pfizer Inc - Dublin (Ireland); Goethe University Frankfurt, University Hospital, Medical Clinic I, Department of Respiratory Medicine, Frankfurt/Main, Germany

**Keywords:** RSV, cardiorespiratory, hospitalization, Germany, quasi-Poisson, time-series modeling

## Abstract

**Introduction:** Respiratory syncytial virus (RSV) can cause severe outcomes among adults. However, RSV incidence in adults is frequently underestimated due to non-specific symptomatology, limited standard-of-care testing, and lower test sensitivity compared to infants. We conducted a retrospective observational study to estimate RSV-attributable incidence of hospitalizations among adults in Germany between 2015 and 2019.

**Methods:** Information on hospitalizations and the number of people at risk of hospitalization (denominator) was gathered from a Statutory Health Insurance (SHI) database. A quasi-Poisson regression model accounting for periodic and aperiodic time trends and virus activity was fitted to estimate the RSV-attributable incidence rate (IR) of four specific cardiovascular hospitalizations (arrhythmia, ischemic heart diseases, chronic heart failure exacerbations, cerebrovascular diseases) and four specific respiratory hospitalizations (influenza/pneumonia, bronchitis/bronchiolitis, chronic lower respiratory tract diseases, upper respiratory tract diseases).

**Results:** RSV-attributable IRs of hospitalizations were generally increasing with age. Among cardiovascular hospitalizations in adults aged ≥60 years, arrhythmia and ischemic heart diseases accounted for the highest incidence of RSV-attributable events, followed by chronic heart failure exacerbation, with annual IR ranges of 157– 260, 133–214, and 105–169 per 100,000 person-years, respectively. The most frequent RSV-attributable respiratory hospitalizations in adults aged ≥60 years were for chronic lower respiratory tract diseases and bronchitis/bronchiolitis, with annual IR ranges of 103–168 and 77–122 per 100,000 person-years, respectively.

**Conclusion:** RSV causes a considerable burden of respiratory and cardiovascular hospitalizations in adults in Germany, similar to other respiratory viruses (e.g., influenza and SARS-CoV-2). This highlights the need to implement effective prevention strategies, especially for older adults.

**Key Summary Points:**

- Respiratory syncytial virus (RSV) disease burden in adults is significant yet often remains unrecognized due to unspecific symptoms, lack of routine testing and lower test sensitivity compared to infants.
- Using a quasi-Poisson regression time-series model, we estimated the age-stratified RSV-attributable incidence of specific cardiovascular and respiratory hospitalizations in Germany between 2015 and 2019.
- Estimated cardiorespiratory RSV hospitalization rates increased with age and were significantly higher in older adults.
- Arrhythmia, ischemic heart diseases, and chronic lower respiratory tract disease exacerbation were the most frequent causes of RSV-attributable cardiovascular and respiratory hospitalizations.
- RSV causes a considerable burden of respiratory and cardiovascular hospitalizations among adults in Germany, and effective RSV vaccines could improve public health outcomes, especially for older adults.

## Introduction

Respiratory syncytial virus (RSV) usually causes mild respiratory infections but may also lead to severe lower respiratory tract diseases such as chronic obstructive pulmonary disease exacerbation and pneumonia (1). RSV affects people of any age, but the risk of severe infections and hospitalizations is highest among infants, adults aged 60 years and above, and those with comorbidities (1–4).

The incidence rate (IR) and clinical burden of RSV disease in adults are challenging to measure due to symptoms resembling influenza and other co-circulating respiratory viruses (5). Limited standard-of-care RSV testing among adults, single specimen collection (i.e., use of nasopharyngeal/nasal swab only) and reduced sensitivity of diagnostic testing compared to children (6–9) contribute to the underestimation of the RSV burden. Additionally, inadequate RSV diagnostic capacity in many healthcare facilities (5, 8, 10, 11) and low awareness of RSV, particularly for patients with cardiopulmonary manifestations, hinder measuring the actual RSV burden (12). Further, even when RSV is diagnosed via laboratory testing, other non-specific acute respiratory infection codes may be used for administrative diagnoses (13).

RSV-associated exacerbation of underlying cardiopulmonary diseases may contribute to a large and unrecognized burden of RSV disease (12). Several studies demonstrated that RSV can cause exacerbation of asthma and chronic obstructive pulmonary disease (COPD) (14–17) (18). Growing evidence indicates that RSV is associated with cardiovascular diseases such as acute myocardial infarction, stroke, arrhythmia, exacerbation of congestive heart failure (CHF), and coronary artery disease (CAD) (12, 19–26). Nonetheless, data on RSV-attributable cardiovascular hospitalizations in adults is limited.

Due to severe under-ascertainment of RSV cases in adults, the recent burden of RSV estimates is more commonly based on mathematical models. Those models relate the temporal variability of the community activity of RSV (indicated by, e.g., week-to-week RSV disease case counts) to the variability in disease outcomes that may be caused or triggered by an RSV infection (e.g., COPD exacerbation hospitalizations and cardiovascular hospitalizations). Among those outcomes, the model estimates the portion expected to be RSV-attributable (10, 27–29).

In Germany, data on the RSV burden from prospective observational incidence studies are unavailable (30), but a recent model-based study showed considerable RSV morbidity and mortality between 2015 and 2019 (31). The estimated RSV-attributable hospitalization IRs increased with age, with high rates observed among people 60 years and above (respiratory diseases: 236–363 per 100,000 person-years; cardiovascular diseases: 558–885 per 100,000 person-years; and cardiorespiratory diseases: 584–912 per 100,000 person-years) (31). However, detailed data on RSV-attributable hospitalizations in more specific respiratory and cardiovascular subgroups are still lacking.

Providing such estimates would help to better understand the burden imposed by RSV and guide the formulation of vaccine policies. Lately, RSV vaccines have been licensed to prevent lower respiratory tract disease caused by RSV in adults 60 years and above. Two RSV vaccines [GSK’s RSV Arexvy vaccine (32) and Pfizer’s RSV Abrysvo vaccine (33)] for adults aged 60 years and above have been available in Germany since 2023. In 2024, market authorization received a third RSV vaccine [Moderna’s mRESVIA vaccine] (34).

We conducted a model-based study to estimate RSV-attributable hospitalizations of four respiratory (influenza/pneumonia, bronchitis/bronchiolitis, chronic lower respiratory tract diseases, upper respiratory tract diseases) and four cardiovascular (chronic heart failure exacerbation, ischemic heart diseases, arrhythmia, cerebrovascular diseases) conditions in Germany.

## Methods

### Study design

We conducted an observational retrospective study to estimate the IRs of specific respiratory and cardiovascular hospitalizations attributable to RSV among adults in Germany using a quasi-Poisson regression model.

### Data sources

Data on hospitalizations was sourced from the Deutsche Analysedatenbank für Evaluation und Versorgungsforschung (DADB) database from Gesundheitsforen Leipzig (GFL). This database contains Statutory Health Insurance (SHI) data of approximately 3.5 million insured individuals. Participants included in the study were adults aged 18 years and above. The age and sex structure of the DADB database is similar to that of the German population, with adjustment factors to account for a slightly younger population in DADB available (35). The study period, from 2015 to 2019, was selected to avoid any distortions in RSV epidemiology due to the COVID-19 pandemic (36).

The study included eight disease outcomes, coded according to the German Modification (GM) of the International Classification of Diseases, 10^th^ revision (ICD-10-GM). These include four specific respiratory outcomes [influenza/ pneumonia (J10-J18), bronchitis/bronchiolitis (J20-J22), chronic lower respiratory tract diseases (J40-J47), upper respiratory tract diseases (J00-J06, J30-J39)], and four specific cardiovascular outcomes [chronic heart failure (I42-I43, I50, I51.7), ischemic heart diseases (I20-I25), arrhythmias (I44-I49), and cerebrovascular diseases (I60-I68)]. To compare the difference between the number of observed (reported in the database) and attributable (model-based) RSV events, we obtained RSV-specific hospitalizations (B97.4, J21.0, J12.1, J20.5) stratified by age group.

Hospitalization was defined as an overnight stay in a hospital commencing from the admission date. If subsequent hospitalizations for the same outcome occurred within 30 days from discharge, these were collapsed with the initial hospitalization to avoid overcounting cases due to readmission.

Individuals were categorized into four age groups: 18-44, 45-59, 60-74, and 75 years and above. The 60-year and above cut-off was selected because RSV vaccines are approved for this age group in Germany. For anonymization purposes, data were suppressed if fewer than 5 cases were reported in the age group and outcome stratum.

The indicator for RSV circulation was defined as the weekly number of RSV-related hospitalizations in children under two years of age (ICD-10-GM codes: B97.4, J21.0, J12.1, J20.5, J21.9). As in other studies (10, 26, 28, 37), we selected RSV circulation in children due to the higher frequency of testing and hospitalization in this age group and the higher sensitivity of diagnostic tests compared to adults (8, 38). As most bronchiolitis cases in young children are caused by RSV, we also included J21.9 (acute bronchiolitis, unspecified) (39–41) as a proxy for RSV circulation. The indicator for influenza circulation was defined as the weekly number of influenza-specific hospitalizations (ICD-10-GM codes: J09, J10, J11) in adults aged 60 years and above (37).

### Statistical analysis

Upon observation of seasonality, age-group-stratified weekly aggregated data were modeled using a quasi-Poisson regression model. The model was constructed according to the previously published protocol (42). The model accounts for baseline periodic and aperiodic time trends, viral activity (RSV and influenza) and potential overdispersion.

With these fitted models, we obtained the annual number of RSV-attributable events and the proportion of RSV-attributable events for each outcome and age group as described in the protocol (42).

Yearly IRs of RSV-attributable events, expressed in the number of events per 100,000 person-years, were obtained by dividing the annual model-based number of RSV-attributable events by the population at risk. This age-specific population at risk of the event was obtained from the DADB database. No effort was made to limit the denominator to persons at risk for the given condition (e.g., estimate COPD exacerbation incidence only among persons with COPD diagnosis). Because the DADB database, from which our estimates were obtained, has a slightly younger population compared to the overall German population covered by SHI, the yearly RSV-attributable events were multiplied with age-specific correction factors based on data provided by GFL to scale the IRs to a nationally representative level.

Confidence intervals were obtained via residual bootstrapping. All data management and statistical analyses were conducted using R software (version 4.0.4).

### Ethical considerations

This study used aggregated and anonymized data, therefore not requiring approval from Institutional Review Boards or Ethical Committees or informed consent from patients. The study was conducted following legal and regulatory requirements and research practices described in the Good Epidemiological Practice guidelines issued by the International Epidemiological Association (43).

## Results

### Observed hospitalizations

Between 2015 and 2019, the diagnoses with the largest number of hospitalizations were due to arrhythmia and ischemic heart diseases, responsible for 266,463 and 259,052 hospitalizations, respectively. Among respiratory hospitalizations, chronic lower respiratory tract diseases were responsible for 162,344 hospitalizations, with influenza/pneumonia (74,064 hospitalizations) accounting for the most (Supplementary Table 1). The majority of hospitalizations, amounting to 70% or more, involved individuals aged 60 years and above. The exception to this trend was observed in upper respiratory tract diseases, where most hospitalizations (74% or more) occurred in younger adults aged 18-59 years.

The reported number of RSV-specific hospitalizations based on ICD code diagnoses alone throughout the study period was 24 cases across all age groups. However, the total number of RSV-specific hospitalizations is unknown as, during most weeks in which RSV-specific hospitalizations were reported, counts were below five and hence suppressed for anonymity.

### Estimated RSV-attributable hospitalizations

The estimated IR of RSV-attributable diseases showed year-to-year fluctuations, with the highest incidence observed for most outcomes in 2017. Due to a lack of adequate seasonal fluctuations, several outcomes were not modeled in the youngest age group. For cerebrovascular diseases, only data for the age group 75 years and above were suitable for modeling (Table 1 and Table 2).

**Table 1.** Estimated incidence rate per 100,000 person-years of RSV-attributable respiratory hospitalizations and percentage (%) of all respective respiratory hospitalizations attributable to RSV infections in adults in Germany, 2015-2019.

| Age groups | 2015 |  | 2016 |  | 2017 |  | 2018 |  | 2019 |  |
| --- | --- | --- | --- | --- | --- | --- | --- | --- | --- | --- |
|  | IR [95% CI] | % | IR [95% CI] | % | IR [95% CI] | % | IR [95% CI] | % | IR [95% CI] | % |
| Influenza/pneumonia |  |  |  |  |  |  |  |  |  |  |
| 18-44 | 2.7<br>[0, 6.0] | 2.9 | 4.2<br>[0, 9.1] | 3.6 | 4.8<br>[0, 10.4] | 4.4 | 3.8<br>[0, 8.4] | 3.1 | 5.0<br>[0, 11.0] | 4.6 |
| 45-59 | 3.3<br>[0, 8.1] | 1.3 | 4.8<br>[0, 11.8] | 1.8 | 5.2<br>[0, 13.0] | 1.9 | 4.1<br>[0, 10.3] | 1.4 | 5.5<br>[0, 13.6] | 2.0 |
| 60-74 | 20.4<br>[0, 42.6] | 1.9 | 20.8<br>[0, 43.6] | 1.8 | 27.7<br>[0, 57.9] | 2.4 | 18.1<br>[0, 37.9] | 1.6 | 23.9<br>[0, 50.0] | 2.2 |
| ≥75 | 231.3<br>[137.4, 330.0] | 4.3 | 253.7<br>[150.7, 362.0] | 4.5 | 292.4<br>[173.7, 417.2] | 5.1 | 205.8<br>[122.3, 293.7] | 3.6 | 273.9<br>[162.7, 390.8] | 5.0 |
| ≥60* | 73.7 | 3.4 | 81.0 | 3.5 | 97.9 | 4.1 | 67 | 2.8 | 88.4 | 3.9 |
| Bronchitis/bronchiolitis |  |  |  |  |  |  |  |  |  |  |
| 18-44 | 0.4<br>[0, 2.7] | 0.9 | 0.5<br>[0, 3.3] | 1.0 | 0.6<br>[0, 4.5] | 1.4 | 0.4<br>[0, 3.2] | 0.9 | 0.6<br>[0, 4.3] | 1.5 |
| 45-59 | 6.6<br>[3.4, 10.1] | 8.6 | 7.7<br>[4.0, 11.9] | 9.2 | 10.0<br>[5.2, 15.4] | 11.0 | 6.9<br>[3.6, 10.7] | 7.8 | 9.7<br>[5.0, 14.9] | 11.1 |
| 60-74 | 25.5<br>[14.9, 35.8] | 9.0 | 24.1<br>[14.1, 33.9] | 8.8 | 34.4<br>[20.1, 48.4] | 11.8 | 21.2<br>[12.4, 29.8] | 7.3 | 28.5<br>[16.7, 40.1] | 10.9 |
| ≥75 | 283.6<br>[222.9, 343.8] | 19.6 | 281.9<br>[221.6, 341.8] | 20.1 | 359.8<br>[282.8, 436.3] | 22.4 | 234.4<br>[184.2, 284.2] | 16.6 | 314.2<br>[247.0, 381.0] | 24.5 |
| ≥60* | 91.3 | 15.5 | 91.5 | 15.9 | 121.8 | 18.7 | 77.2 | 13.1 | 102.9 | 19.3 |
| Chronic lower respiratory tract diseases |  |  |  |  |  |  |  |  |  |  |
| 18-44 | NA |  | NA |  | NA |  | NA |  | NA |  |
| 45-59 | 18.9<br>[1.7, 35.5] | 2.4 | 17.4<br>[1.5, 32.7] | 2.0 | 28.1<br>[2.5, 52.8] | 3.1 | 17.5<br>[1.6, 32.9] | 2.0 | 25.4<br>[2.3, 47.8] | 2.9 |
| 60-74 | 74.4<br>[17.2, 133.8] | 2.4 | 70.4<br>[16.3, 126.6] | 2.1 | 100.5<br>[23.2, 180.7] | 3.0 | 61.8<br>[14.3, 111.2] | 1.9 | 83.3<br>[19.2, 149.8] | 2.6 |
| ≥75 | 279.5<br>[125.7, 432.5] | 3.3 | 239.0<br>[107.5, 369.8] | 2.6 | 342.7<br>[154.1, 530.3] | 3.9 | 211.4<br>[95.1, 327.1] | 2.6 | 293.0<br>[131.7, 453.3] | 3.6 |
| ≥60* | 128.9 | 2.8 | 116.7 | 2.3 | 168.2 | 3.4 | 102.9 | 2.2 | 140.1 | 3.1 |
| Upper respiratory tract diseases |  |  |  |  |  |  |  |  |  |  |
| 18-44 | 10.9<br>[0, 27.3] | 1.7 | 10.4<br>[0, 26.1] | 1.5 | 17.4<br>[0, 43.7] | 2.4 | 11.1<br>[0, 27.8] | 1.6 | 15.8<br>[0, 39.7] | 2.3 |
|  | IR [95% CI] | % | IR [95% CI] | % | IR [95% CI] | % | IR [95% CI] | % | IR [95% CI] | % |
| 45-59 | 8.2<br>[0, 21.7] | 1.9 | 8.1<br>[0, 21.3] | 1.7 | 12.4<br>[0, 32.9] | 2.5 | 8.0<br>[0, 21.1] | 1.7 | 11.4<br>[0, 30.1] | 2.4 |
| 60-74 | 18.6<br>[0.4, 35.5] | 3.6 | 17.6<br>[0.4, 33.6] | 3.1 | 25.1<br>[0.6, 48.0] | 4.4 | 15.4<br>[0.4, 29.5] | 2.8 | 20.8<br>[0.5, 39.8] | 3.9 |
| ≥75 | 55.9<br>[25.7, 86.4] | 7.2 | 51.2<br>[23.5, 79.1] | 6.4 | 70.0<br>[32.2, 108.2] | 8.4 | 44.5<br>[20.4, 68.7] | 5.5 | 60.6<br>[27.8, 93.5] | 8.1 |
| ≥60* | 28.7 | 4.8 | 26.9 | 4.1 | 37.9 | 5.8 | 23.5 | 3.7 | 31.7 | 5.2 |

*CI: confidence interval; IR: incidence rate extrapolated to the national level; NA: not applicable due to data not suitable for modeling*
*\* Based on pooling results for models fitted to adults for included age groups, thus confidence intervals are not provided.*

**Table 2.**
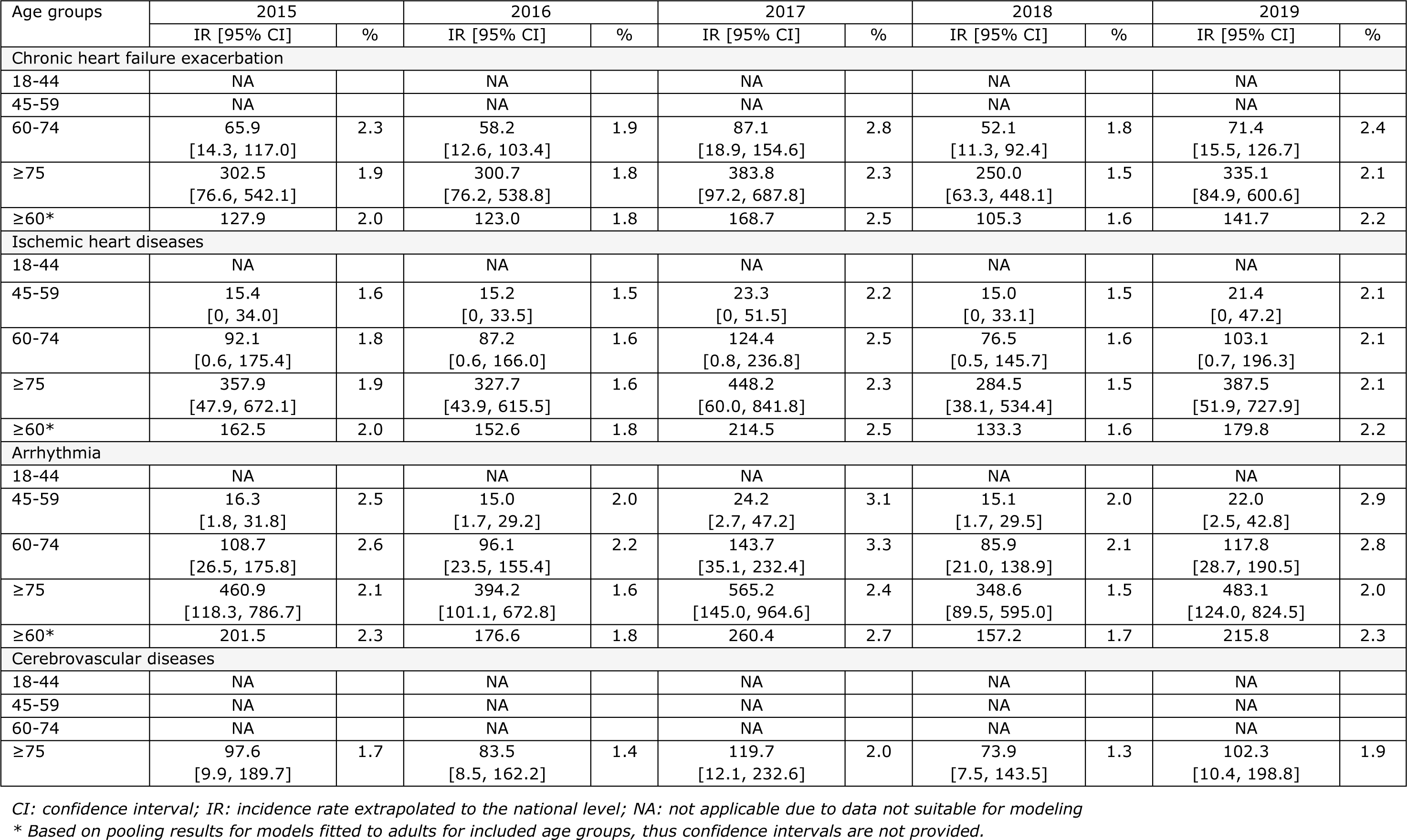
Estimated incidence rate per 100,000 person-years of RSV-attributable cardiovascular hospitalizations and percentage (%) of all respective cardiovascular hospitalizations attributable to RSV infections in adults in Germany, 2015-2019.

The estimated IR of RSV-attributable respiratory hospitalizations in the age group 60 years and above was highest for chronic lower respiratory tract diseases (range across the study period: 103-168 hospitalizations per 100,000) and bronchitis/ bronchiolitis (77-122 hospitalizations per 100,000) (Table 1). The IRs of RSV-attributable bronchitis/bronchiolitis hospitalizations for those aged 75 years and above were approximately 11- to 12-fold and 32- to 43-fold higher than the rates for adults aged 60-74 years and 45-59 years, respectively. Similarly, the rates of chronic lower respiratory diseases were approximately 3- to 4-fold and 12- to 15- fold higher in the oldest age group (≥75 years) compared to younger age groups (60-74 and 45-59 years) (Table 1).

RSV-attributable proportions were comparable between age groups and respiratory subgroups (range 1-5% in age groups 45-59 years and 60-74 years, and 3-8% in ≥75 years) with the exception of bronchitis or bronchiolitis where the RSV-attributable proportion was 1-10% for younger age groups and 17-25% for adults 75 years and above (Table 1). For cardiovascular causes, arrhythmia (157-260 per 100,000 person-years) and ischemic heart diseases (133-215 per 100,000 person-years), followed by chronic heart failure (105-169 per 100,000 person-years) showed the highest RSV-attributable IRs (Table 2) in adults aged ≥60 years. The RSV-attributable IRs were notably higher in older age groups, particularly among hospitalized individuals aged 75 years and above. The rates of RSV-attributable arrhythmia and ischemic heart disease hospitalizations for those aged 75 and above were both approximately 4-fold higher than the rates for adults aged 60-74 years. Moreover, these rates were about 25-fold and 21-fold higher, respectively, than for adults aged 45-59 years (Table 2). RSV-attributable proportions were comparable between age groups and cardiovascular subgroups, accounting for approximately 1% to 3% of all hospitalizations (Table 2).

## Discussion

Our study complements the results of the first model-based study in Germany (31) by providing estimates on specific types of cardiorespiratory hospitalizations attributable to RSV. We found a high burden of cardiovascular hospitalizations related to RSV, with RSV-attributable IRs for arrhythmia (157-260 per 100,000 person-years) and ischemic heart diseases (133-215 per 100,000 person-years) being the two highest among all studied conditions in adults aged 60 years and above. Among respiratory conditions, chronic lower respiratory tract diseases (103-168 per 100,000 person-years) and bronchitis/bronchiolitis (77-122 per 100,000 person-years) had the highest RSV-attributable incidences in adults aged 60 years and over. As in other studies, we found that RSV-attributable IRs were increasing with age, with a considerable surge observed in patients aged 60 years and older. Our findings were considerably higher than those based on RSV-specific ICD-10 codes alone, suggesting vast under-ascertainment of RSV cases in adults. Substantially lower RSV hospitalization rates in adults aged >59 years (0.11-11.36 per 100,000 person-years) were also reported in an observational study in Germany, which was based on data on primary discharge diagnosis of RSV (J21.1, J20.5, J21.0) (44).

Our study adds to the growing evidence of the association between RSV circulation and cardiovascular events. We found that approximately 2% to 3% of hospitalizations due to arrhythmia, ischemic heart diseases, and chronic heart failure in people 60 years and above were attributed to RSV. In the oldest age group (75 years and above), we also found an association – in line with another study – between RSV and cerebrovascular diseases, with 1% to 2% of those hospitalizations being attributable to RSV. For instance, in Spain, the RSV-attributable proportion of ischemic heart diseases in adults aged 60 years and above (3%) and cerebrovascular diseases in adults aged 80 years and above (2%) were comparable to our results (26). Although the RSV-attributable proportions were comparable, it resulted in higher IRs, most likely due to overall higher observed hospitalizations due to cardiovascular causes in Germany. This might be related to different coding practices or different prevalence of comorbidities in those two populations.

Several mechanisms are considered to explain the relationship between RSV and cardiovascular events. Part of those conditions may be related to the indirect effect of inflammation due to acute respiratory infections. Inflammation may increase the concentration of C-reactive protein, inflammatory cytokines and clotting factors such as fibrinogen, leading to an increased risk of thrombotic coronary occlusion (20, 45, 46). Additionally, it may cause endothelial dysfunction and inhibit the function of vasodilating nitric oxide or prostaglandins, contributing to arterial and venous thromboembolic disease (47). Development of pulmonary hypertension may lead to secondary myocardial dysfunction, hypotension and arrhythmia (24). RSV can also directly infect organs, as it was isolated from myocardial tissue biopsies in patients with myocarditis (19, 24). Cardiovascular diseases remain the leading cause of morbidity and mortality worldwide (48).

Thus, the potential impact of a vaccine preventing cardiovascular events, as observed for the influenza vaccine (49, 50), might be substantial. Our estimates for RSV-attributable hospitalizations due to bronchitis/ bronchiolitis (77-122 per 100,000) and influenza/pneumonia diagnosis grouping (67-98 per 100,000) in adults aged 60 years and above align with the study conducted in Spain (100-110 per 100,000 and 83-91 and, respectively) (26). The Spanish study utilized the same generic protocol that we used for this study. In contrast, when compared to another model-based study from the United Kingdom, IRs in older age groups (65-74 and ≥75 years) were considerably higher in Germany (10). The possible reason is using different methods (quasi-Poisson regression modeling weekly counts vs linear regression modeling weekly outcome rates) and a different RSV proxy definition (weekly hospitalizations due to RSV vs weekly RSV counts from the surveillance system). We used RSV hospitalization data as a proxy because hospitalization data is less affected by the testing practices than the surveillance data, which will be mostly driven by influenza (data not shown). Further, the UK study used primary diagnosis codes, which have been shown to underestimate incidence in validation studies (51).

Among age groups 45-59 and 60-74 years, the highest RSV-attributable IR was revealed for chronic lower respiratory tract diseases. Respiratory viruses, including RSV, are known causes of exacerbation of COPD (16). In prospective cohort studies, approximately 4% to 11% of COPD patients tested positive for RSV (52, 53). The prevalence of COPD is increasing globally, leading to enormous healthcare costs and high mortality (54, 55). Exacerbation of COPD is associated with an accelerated decline in lung function, progression of the disease and lower quality of life (15). Thus, preventing COPD exacerbation should be a public health priority, and vaccination of COPD patients should be prioritized, as suggested by the recent recommendation from the US Centers for Disease Control and Prevention (56).

Our study showed considerable under-ascertainment of RSV cases in adults in Germany. Other studies also observed low testing activity and considerable underestimation of the RSV burden in adults (8, 9, 26) when only RSV-specific ICD codes are used. These findings highlight the need for systematic testing and reporting, including patients presenting with cardiovascular manifestations or exacerbation of underlying respiratory diseases.

The main strength of our study was the use of a large database of approximately 3.5 million insured persons, an extended study period of five years and the inclusion of eight different medical conditions, allowing for a more sensitive estimation of the RSV burden. In addition, our study was based on a generic protocol (42) that permits robust comparisons between countries. The protocol was developed based on an extensive prior literature review and experts’ input.

We also acknowledge some study limitations. Our model included only RSV and influenza as viral indicators, assuming those are the only pathogens related to the outcome of interest. However, by inclusion of the periodic component and overdispersion parameter, we indirectly accounted for other potentially relevant pathogens. Second, RSV hospitalizations in children less than two years old as the RSV indicator may not fully reflect viral activity in adults. However, the pattern in adults is similar to that observed in children, and for the model performance, the most important factor is the fluctuation over time rather than the absolute numbers. Third, although the SHI database is representative of the German population, the age distribution is slightly skewed towards young individuals. To account for this difference, we used correction factors to standardize for age in the incidence calculation. Lastly, we did not limit the population denominators to those individuals at risk of a specific condition (e.g., limit the denominator for the RSV-attributable COPD events to those with COPD diagnosis), which would likely increase the reported incidence rate.

In conclusion, our study showed a high and largely unrecognized cardiovascular and respiratory burden of RSV among adults in Germany. Substantial underestimation of RSV cases emphasizes the need for standard-of-care testing, also among adults with cardiovascular symptoms. Newly introduced vaccines might have a high impact, not only on respiratory hospitalization but also on cardiac hospitalizations.

## Medical Writing, Editorial, and Other Assistance

Editorial assistance in the preparation of this article was provided by Dr. Somsuvro Basu of P95. Support for this assistance was funded by Pfizer Ltd.

## Data availability

The datasets generated and/or analyzed during this study are not publicly available (data provider’s restrictions). Weekly counts of hospitalized cases and extrapolation factors were provided by the Gesundheitsforen Leipzig GmbH.

## References

1. Falsey AR. Respiratory syncytial virus infection in adults. Semin Respir Crit Care Med 2007;28:171–81.

2. Falsey AR, Hennessey PA, Formica MA, Cox C, Walsh EE. Respiratory syncytial virus infection in elderly and high-risk adults. N Engl J Med 2005;352:1749–59.

3. Falsey AR. Respiratory Syncytial Virus Infection in Elderly Adults. Drugs Aging 2005;22:578–87.

4. Njue A, Nuabor W, Lyall M, Margulis A, Mauskopf J, Curcio D, et al. Systematic Literature Review of Risk Factors for Poor Outcomes Among Adults With RSV Infection in High-Income Countries. Open Forum Infect Dis 2023;ofad513.

5. Rozenbaum MH, Begier E, Kurosky SK, Whelan J, Bem D, Pouwels KB, et al. Incidence of Respiratory Syncytial Virus Infection in Older Adults: Limitations of Current Data. Infect Dis Ther 2023;12:1487–504.

6. Ramirez J, Carrico R, Wilde A, Junkins A, Furmanek S, Chandler T, et al. Diagnosis of Respiratory Syncytial Virus in Adults Substantially Increases When Adding Sputum, Saliva, and Serology Testing to Nasopharyngeal Swab RT-PCR. Infect Dis Ther 2023;12:1593–603.

7. Onwuchekwa C, Moreo LM, Menon S, Machado B, Curcio D, Kalina W, et al. Underascertainment of Respiratory Syncytial Virus Infection in Adults Due to Diagnostic Testing Limitations: A Systematic Literature Review and Meta-analysis. J Infect Dis 2023;228:173–84.

8. Rozenbaum MH, Judy J, Tran D, Yacisin K, Kurosky SK, Begier E. Low Levels of RSV Testing Among Adults Hospitalized for Lower Respiratory Tract Infection in the United States. Infect Dis Ther 2023;12:677–85.

9. McLaughlin JM, Khan F, Begier E, Swerdlow DL, Jodar L, Falsey AR. Rates of Medically Attended RSV Among US Adults: A Systematic Review and Meta-analysis. Open Forum Infect Dis 2022;9:ofac300.

10. Fleming DM, Taylor RJ, Lustig RL, Schuck-Paim C, Haguinet F, Webb DJ, et al. Modelling estimates of the burden of Respiratory Syncytial virus infection in adults and the elderly in the United Kingdom. BMC Infect Dis 2015;15:443.

11. Ambrosch A, Luber D, Klawonn F, Kabesch M. Focusing on severe infections with the respiratory syncytial virus (RSV) in adults: Risk factors, symptomatology and clinical course compared to influenza A / B and the original SARS-CoV-2 strain. J Clin Virol 2023;161:105399.

12. Branche AR, Saiman L, Walsh EE, Falsey AR, Sieling WD, Greendyke W, et al. Incidence of Respiratory Syncytial Virus Infection Among Hospitalized Adults, 2017-2020. Clin Infect Dis 2022;74:1004–11.

13. Cai W, Tolksdorf K, Hirve S, Schuler E, Zhang W, Haas W, et al. Evaluation of using ICD-10 code data for respiratory syncytial virus surveillance. Influenza Other Respir Viruses 2020;14:630–7.

14. Adeli M, El-Shareif T, Hendaus MA. Asthma exacerbation related to viral infections: An up to date summary. J Family Med Prim Care 2019;8:2753–9.

15. Linden D, Guo-Parke H, Coyle PV, Fairley D, McAuley DF, Taggart CC, et al. Respiratory viral infection: a potential “missing link” in the pathogenesis of COPD. Eur Respir Rev 2019;28.

16. Rohde G, Wiethege A, Borg I, Kauth M, Bauer TT, Gillissen A, et al. Respiratory viruses in exacerbations of chronic obstructive pulmonary disease requiring hospitalisation: a case-control study. Thorax 2003;58:37–42.

17. Ramaswamy M, Groskreutz DJ, Look DC. Recognizing the importance of respiratory syncytial virus in chronic obstructive pulmonary disease. Copd 2009;6:64–75.

18. Westerly BD, Peebles RS, Jr. Respiratory syncytial virus infections in the adult asthmatic--mechanisms of host susceptibility and viral subversion. Immunol Allergy Clin North Am 2010;30:523–39, vi–vii.

19. Ivey KS, Edwards KM, Talbot HK. Respiratory Syncytial Virus and Associations With Cardiovascular Disease in Adults. J Am Coll Cardiol 2018;71:1574–83.

20. Bosco E, van Aalst R, McConeghy KW, Silva J, Moyo P, Eliot MN, et al. Estimated Cardiorespiratory Hospitalizations Attributable to Influenza and Respiratory Syncytial Virus Among Long-term Care Facility Residents. JAMA network open 2021;4:e2111806.

21. Kawashima H, Inagaki N, Nakayama T, Morichi S, Nishimata S, Yamanaka G, et al. Cardiac Complications Caused by Respiratory Syncytial Virus Infection: Questionnaire Survey and a Literature Review. Glob Pediatr Health 2021;8:2333794×211044114.

22. Nguyen JL, Yang W, Ito K, Matte TD, Shaman J, Kinney PL. Seasonal Influenza Infections and Cardiovascular Disease Mortality. JAMA Cardiol 2016;1:274–81.

23. Eisenhut M. Extrapulmonary manifestations of severe respiratory syncytial virus infection--a systematic review. Crit Care 2006;10:R107.

24. Gkentzi D, Dimitriou G, Karatza A. Non-pulmonary manifestations of respiratory syncytial virus infection. J Thorac Dis 2018;10:S3815–s8.

25. Franczuk P, Tkaczyszyn M, Kulak M, Domenico E, Ponikowski P, Jankowska EA. Cardiovascular Complications of Viral Respiratory Infections and COVID-19. Biomedicines 2022;11.

26. Haeberer M, Bruyndonckx R, Polkowska-Kramek A, Torres A, Liang C, Nuttens C, et al. Estimated Respiratory Syncytial Virus-Related Hospitalizations and Deaths Among Children and Adults in Spain, 2016-2019. Infect Dis Ther 2024.

27. Matias G, Taylor RJ, Haguinet F, Schuck-Paim C, Lustig RL, Shinde V. Estimates of mortality attributable to influenza and RSV in the United States during 1997-2009 by influenza type or subtype, age, cause of death, and risk status. Influenza and other respiratory viruses 2014;8:507–15.

28. Goldstein E, Greene SK, Olson DR, Hanage WP, Lipsitch M. Estimating the hospitalization burden associated with influenza and respiratory syncytial virus in New York City, 2003-2011. Influenza Other Respir Viruses 2015;9:225–33.

29. Zhou H, Thompson WW, Viboud CG, Ringholz CM, Cheng PY, Steiner C, et al. Hospitalizations associated with influenza and respiratory syncytial virus in the United States, 1993-2008. Clinical infectious diseases : an official publication of the Infectious Diseases Society of America 2012;54:1427–36.

30. Li Y, Kulkarni D, Begier E, Wahi-Singh P, Wahi-Singh B, Gessner B, et al. Adjusting for Case Under-Ascertainment in Estimating RSV Hospitalisation Burden of Older Adults in High-Income Countries: a Systematic Review and Modelling Study. Infect Dis Ther 2023;12:1137–49.

31. Polkowska-Kramek A, Begier E, Bruyndonckx R, Liang C, Beese C, Brestrich G, et al. Estimated Incidence of Hospitalizations and Deaths Attributable to Respiratory Syncytial Virus Infections Among Adults in Germany Between 2015 and 2019. Infect Dis Ther 2024.

32. Papi A, Ison MG, Langley JM, Lee DG, Leroux-Roels I, Martinon-Torres F, et al. Respiratory Syncytial Virus Prefusion F Protein Vaccine in Older Adults. N Engl J Med 2023;388:595–608.

33. Walsh EE, Pérez Marc G, Zareba AM, Falsey AR, Jiang Q, Patton M, et al. Efficacy and Safety of a Bivalent RSV Prefusion F Vaccine in Older Adults. N Engl J Med 2023;388:1465–77.

34. Moderna Receives U.S. FDA Approval for RSV Vaccine mRESVIA(R). https://investors.modernatx.com/news/news-details/2024/Moderna-Receives-U.S.-FDA-Approval-for-RSV-Vaccine-mRESVIAR/default.aspx.

35. GmbH GL. Deutsche Analysedatenbank für Evaluation und Versorgungsforschung, Stand Januar. 2022. https://www.gesundheitsforen.net/GFL/Services/Analytik/Gesundheitsforen%20Leipzig_DADB_Q1%202024.pdf. Date last accessed: 19.04.2024 2024.

36. Oh DY, Buda S, Biere B, Reiche J, Schlosser F, Duwe S, et al. Trends in respiratory virus circulation following COVID-19-targeted nonpharmaceutical interventions in Germany, January - September 2020: Analysis of national surveillance data. Lancet Reg Health Eur 2021;6:100112.

37. Zheng Z, Warren JL, Shapiro ED, Pitzer VE, Weinberger DM. Estimated incidence of respiratory hospitalizations attributable to RSV infections across age and socioeconomic groups. Pneumonia (Nathan) 2022;14:6.

38. Casiano-Colón AE, Hulbert BB, Mayer TK, Walsh EE, Falsey AR. Lack of sensitivity of rapid antigen tests for the diagnosis of respiratory syncytial virus infection in adults. J Clin Virol 2003;28:169–74.

39. Calvo C, Pozo F, García-García ML, Sanchez M, Lopez-Valero M, Pérez-Breña P, et al. Detection of new respiratory viruses in hospitalized infants with bronchiolitis: a three-year prospective study. Acta Paediatr 2010;99:883–7.

40. Mansbach JM, Piedra PA, Teach SJ, Sullivan AF, Forgey T, Clark S, et al. Prospective multicenter study of viral etiology and hospital length of stay in children with severe bronchiolitis. Arch Pediatr Adolesc Med 2012;166:700–6.

41. Kenmoe S, Kengne-Nde C, Ebogo-Belobo JT, Mbaga DS, Fatawou Modiyinji A, Njouom R. Systematic review and meta-analysis of the prevalence of common respiratory viruses in children < 2 years with bronchiolitis in the pre-COVID-19 pandemic era. PLoS One 2020;15:e0242302.

42. Bruyndonckx R, Polkowska-Kramek A, Liang C, Nuttens C, Tran TMP, Gessner BD, et al. Estimation of Symptomatic Respiratory Syncytial Virus Infection Incidence in Adults in Multiple Countries: A Time-Series Model-Based Analysis Protocol. Infect Dis Ther 2024.

43. Good Epidemiological Practice. Guidelines for Proper Conduct of Epidemiological Research, (2007).

44. Niekler P, Goettler D, Liese JG, Streng A. Hospitalizations due to respiratory syncytial virus (RSV) infections in Germany: a nationwide clinical and direct cost data analysis (2010-2019). Infection 2023.

45. Meier CR, Jick SS, Derby LE, Vasilakis C, Jick H. Acute respiratory-tract infections and risk of first-time acute myocardial infarction. Lancet 1998;351:1467–71.

46. Smeeth L, Thomas SL, Hall AJ, Hubbard R, Farrington P, Vallance P. Risk of myocardial infarction and stroke after acute infection or vaccination. N Engl J Med 2004;351:2611–8.

47. Vallance P, Collier J, Bhagat K. Infection, inflammation, and infarction: does acute endothelial dysfunction provide a link? Lancet 1997;349:1391–2.

48. Roth GA, Mensah GA, Johnson CO, Addolorato G, Ammirati E, Baddour LM, et al. Global Burden of Cardiovascular Diseases and Risk Factors, 1990-2019: Update From the GBD 2019 Study. J Am Coll Cardiol 2020;76:2982–3021.

49. Zangiabadian M, Nejadghaderi SA, Mirsaeidi M, Hajikhani B, Goudarzi M, Goudarzi H, et al. Protective effect of influenza vaccination on cardiovascular diseases: a systematic review and meta-analysis. Sci Rep 2020;10:20656.

50. Nichol KL, Nordin J, Mullooly J, Lask R, Fillbrandt K, Iwane M. Influenza vaccination and reduction in hospitalizations for cardiac disease and stroke among the elderly. N Engl J Med 2003;348:1322–32.

51. Hanquet G, Theilacker C, Vietri J, Sepúlveda-Pachón I, Menon S, Gessner B, et al. Best Practices for Identifying Hospitalized Lower Respiratory Tract Infections Using Administrative Data: A Systematic Literature Review of Validation Studies. Infect Dis Ther 2024;13:921–40.

52. Wiseman DJ, Thwaites RS, Ritchie AI, Finney L, Macleod M, Kamal F, et al. RSV-related Community COPD Exacerbations and Novel Diagnostics: A Binational Prospective Cohort Study. Am J Respir Crit Care Med 2024.

53. Colosia AD, Yang J, Hillson E, Mauskopf J, Copley-Merriman C, Shinde V, et al. The epidemiology of medically attended respiratory syncytial virus in older adults in the United States: A systematic review. PLoS One 2017;12:e0182321.

54. Agarwal D. COPD generates substantial cost for health systems. Lancet Glob Health 2023;11:e1138–e9.

55. Chen S, Kuhn M, Prettner K, Yu F, Yang T, Bärnighausen T, et al. The global economic burden of chronic obstructive pulmonary disease for 204 countries and territories in 2020-50: a health-augmented macroeconomic modelling study. Lancet Glob Health 2023;11:e1183–e93.

56. Melgar M, Britton A, Roper LE, Talbot HK, Long SS, Kotton CN, et al. Use of Respiratory Syncytial Virus Vaccines in Older Adults: Recommendations of the Advisory Committee on Immunization Practices - United States, 2023. MMWR Morb Mortal Wkly Rep 2023;72:793–801.

