## Supplementary file for "Estimated incidence rate of specific cardiovascular and respiratory hospitalizations attributable to Respiratory Syncytial Virus among adults in Germany between 2015 and 2019"


### Author Details:

Caihua Liang[1], Aleksandra Polkowska-Kramek[2], Caroline Lade[3], Lea Johanna Bayer[3], Robin Bruyndonckx[2], Bennet Huebbe[3], Worku Biyadgie Ewnetu[2], Pimnara Peerawaranun[2], Maribel Casas[2], Thao Mai Phuong Tran[2], Gordon Brestrich[3], Christof von Eiff[3], Bradford D. Gessner[1], Elizabeth Begier[4], Gernot Rohde[5]

### Affiliations:

[1] Pfizer Inc-New York (USA)

[2] P95 Epidemiology & Pharmacovigilance, Leuven, Belgium

[3] Pfizer Pharma GmbH - Berlin (Germany)

[4] Pfizer Inc - Dublin (Ireland)

[5] Goethe University Frankfurt, University Hospital, Medical Clinic I, Department of Respiratory Medicine, Frankfurt/Main, Germany

### Corresponding author:

Caihua Liang

Address: 66 Hudson Blvd E, New York, NY 10001

### Supplementary Table 1. Number of hospitalized cases stratified by age group in Germany, 2015-2019

| | Age group | | | | |
|---|---|---|---|---|---|
| Outcome | 18-44 years | 45-59 years | 60-74 years | ≥75 years | Total |
| **Influenza/pneumonia** | | | | | |
| **2015*** | 848 | 1893 | 3557 | 5745 | 12043 |
| **2016** | 1043 | 2149 | 4035 | 6527 | 13754 |
| **2017** | 957 | 2258 | 4550 | 7733 | 15498 |
| **2018** | 1090 | 2498 | 4860 | 8112 | 16560 |
| **2019** | 959 | 2277 | 4948 | 8025 | 16209 |
| **Total** | 4897 | 11075 | 21950 | 36142 | 74064 |
| **Bronchitis/bronchiolitis** | | | | | |
| **2015*** | 360 | 561 | 925 | 1499 | 3345 |
| **2016** | 374 | 631 | 947 | 1584 | 3536 |
| **2017** | 358 | 706 | 1116 | 2105 | 4285 |
| **2018** | 414 | 737 | 1233 | 1995 | 4379 |
| **2019** | 346 | 707 | 1162 | 1875 | 4090 |
| **Total** | 1852 | 3342 | 5383 | 9058 | 19635 |
| **Chronic lower respiratory tract diseases** | | | | | |
| **2015*** | 2319 | 5918 | 10069 | 8722 | 27028 |
| **2016** | 2506 | 6529 | 11440 | 10380 | 30855 |
| **2017** | 2613 | 6936 | 12608 | 11626 | 33783 |
| **2018** | 2668 | 7054 | 13517 | 11630 | 34869 |
| **2019** | 2572 | 7130 | 14102 | 12005 | 35809 |
| **Total** | 12678 | 33567 | 61736 | 54363 | 162344 |
| **Upper respiratory tract diseases** | | | | | |
| **2015*** | 5453 | 3228 | 1668 | 804 | 11153 |
| **2016** | 5892 | 3589 | 1952 | 911 | 12344 |
| **2017** | 5974 | 3780 | 2154 | 1086 | 12994 |
| **2018** | 6107 | 3853 | 2315 | 1141 | 13416 |
| **2019** | 5907 | 3819 | 2347 | 1098 | 13171 |
| **Total** | 29333 | 18269 | 10436 | 5040 | 63078 |
| **Chronic heart failure exacerbation** | | | | | |
| **2015*** | 592 | 3744 | 9333 | 16411 | 30080 |
| **2016** | 673 | 4094 | 10595 | 19283 | 34645 |
| **2017** | 728 | 4483 | 11653 | 21816 | 38680 |
| **2018** | 710 | 4603 | 12175 | 22957 | 40445 |
| **2019** | 715 | 4550 | 13027 | 23777 | 42069 |

| | Age group | | | | |
|---|---|---|---|---|---|
| **Outcome** | **18-44 years** | **45-59 years** | **60-74 years** | **≥75 years** | **Total** |
| **Total** | 3418 | 21474 | 56783 | 104244 | 185919 |
| **Ischemic heart diseases** | | | | | |
| **2015*** | 694 | 7104 | 16212 | 19442 | 43452 |
| **2016** | 722 | 7846 | 18213 | 22615 | 49396 |
| **2017** | 729 | 8161 | 19209 | 25073 | 53172 |
| **2018** | 768 | 8446 | 20014 | 26228 | 55456 |
| **2019** | 800 | 8466 | 21432 | 26878 | 57576 |
| **Total** | 3713 | 40023 | 95080 | 120236 | 259052 |
| **Arrhythmia** | | | | | |
| **2015*** | 1370 | 4826 | 13433 | 22842 | 42471 |
| **2016** | 1522 | 5741 | 15257 | 27226 | 49746 |
| **2017** | 1575 | 5953 | 16563 | 31096 | 55187 |
| **2018** | 1642 | 6231 | 17298 | 32930 | 58101 |
| **2019** | 1595 | 6239 | 18472 | 34652 | 60958 |
| **Total** | 7704 | 28990 | 81023 | 148746 | 266463 |
| **Cerebrovascular diseases** | | | | | |
| **2015*** | 427 | 1870 | 3974 | 6066 | 12337 |
| **2016** | 423 | 2086 | 4380 | 6949 | 13838 |
| **2017** | 462 | 2164 | 4537 | 7760 | 14923 |
| **2018** | 479 | 2291 | 4849 | 8024 | 15643 |
| **2019** | 462 | 2166 | 5042 | 8013 | 15683 |
| **Total** | 2253 | 10577 | 22782 | 36812 | 72424 |

*Weeks 1-5 are not included in the analysis as insufficient information on the RSV proxy is available for these first weeks.
